# Deep brain stimulation modulates directional limbic connectivity in major depressive disorder

**DOI:** 10.1101/2023.05.18.23290168

**Authors:** Egill A. Fridgeirsson, I.O. Bergfeld, B.P. de Kwaasteniet, J. Luigjes, J. van Laarhoven, P. Notten, G. Beute, P. van den Munckhof, P.R. Schuurman, D.A.J.P. Denys, G.A. van Wingen

## Abstract

Deep brain stimulation (DBS) is being investigated as treatment for patients with refractory major depressive disorder (MDD). However, little is known about how DBS exerts its antidepressive effects. Here, we investigated whether ventral anterior limb of the internal capsule (vALIC) stimulation modulates a limbic network centered around the amygdala in patients with treatment resistant MDD.

Nine patients underwent resting state functional magnetic resonance imaging (fMRI) before DBS surgery and after one year of treatment. In addition, they were scanned twice within two weeks during the subsequent double blind crossover phase with active and sham treatment. Eleven matched controls underwent fMRI scans at same time intervals to account for test-retest effects. The imaging data was investigated with functional connectivity analysis and dynamic causal modelling (DCM).

Results showed that one year of DBS treatment was associated with increased functional connectivity of the left amygdala with precentral cortex and left insula along with decreased bilateral connectivity between nucleus accumbens and ventromedial prefrontal cortex. No changes in functional connectivity were observed during the crossover phase. Effective connectivity analyses using DCM revealed widespread amygdala-centric changes between pre-surgery and one year follow-up, while the crossover phase was associated with insula-centric changes between active and sham stimulation.

These results suggest that vALIC DBS results in complex rebalancing of the limbic network involved in emotion, reward and interoceptive processing.

## Introduction

Major depressive disorder (MDD) is a debilitating psychiatric disorder that is one of the leading causes of chronic disease burden in the world (Ferrari et al., 2013). Standard treatments for patients with MDD include pharmacotherapy, psychotherapy and electroconvulsive therapy. An investigational treatment option for patients that do not respond to any of these treatments is deep brain stimulation (DBS), which has shown efficacy of around 40% (Bergfeld et al., 2016a; Bergfeld & Figee, 2020a). In DBS, electrodes are inserted deep into certain brain regions which can then be stimulated with electrical current. The first brain target for DBS in depression was the white matter of the subcallosal cingulate (Mayberg et al., 2005). Subsequently different targets have been explored such as the ventral capsule/ventral striatum (Dougherty et al., 2015; Malone et al., 2009), the nucleus accumbens (NAc) (Schlaepfer et al., 2008a), the superolateral branch of the medial forebrain bundle (Schlaepfer et al., 2013) and the ventral anterior limb of the internal capsule (vALIC) (Bergfeld et al., 2016b; Van Der Wal et al., 2020).

The mechanism for how DBS exert its antidepressant effects is largely unknown. Mood disorders have been consistently associated with increased activity of amygdala and insula, and decreased activity of the prefrontal cortex and nucleus accumbens (NAc) (Etkin & Wager, 2007; J. P. Hamilton et al., 2012; Simon et al., 2014; Taylor & Whalen, 2015; Via et al., 2014; Yang et al., 2022). We recently found that long-term vALIC DBS in MDD normalizes amygdala responsivity to emotionally salient stimuli. In addition, active compared to sham stimulation increased amygdala connectivity with sensorimotor and cingulate cortices (Runia et al., n.d.). Other studies investigating the neural effects of DBS have used the NAc as a target, which is located just below the vALIC target (Bergfeld & Figee, 2020b). Some positron emission tomography studies suggest widespread effects of NAc stimulation on brain metabolism such as decreases in posterior cingulate, caudate nucleus, thalamus, cerebellum and dorsomedial prefrontal gyrus and increases in ventral striatum, dorsolateral and medial prefrontal cortices (Bewernick et al., 2010; Millet et al., 2014; Schlaepfer et al., 2008b). However, it remains unknown how DBS affects the interaction between brain networks that are thought to be at the core of depression. We previously investigated how vALIC DBS affects resting state functional connectivity between the amygdala, the insula, the NAc and the ventromedial prefrontal cortex (vmPFC) in OCD (Fridgeirsson et al., 2020a), which showed how improvements in mood with DBS were associated with reduced amygdala-insula functional connectivity and that DBS decreased directional impact of vmPFC on amygdala and amygdala on the insula. To investigate whether vALIC DBS has comparable effects in MDD, we here investigated the same network.

We used two methods to explore this network using resting state functional magnetic resonance imaging (fMRI). Following our previous studies in OCD, we used seed based functional connectivity analyses using the amygdala and NAc as seeds (Figee et al., 2013a; Fridgeirsson et al., 2020b). Then we used an effective connectivity analysis based on spectral dynamic causal modelling (K. J. Friston et al., 2014a).

This type of analysis allow us to estimate directional connectivity and excitation/inhibition balance of regions. We tested whether there are connectivity changes from before the DBS surgery and treatment initiation to after the DBS parameters have been optimized to maximize treatment response. We controlled for test-retest effects by measuring healthy controls around the same time points. And we then tested whether there are short term changes during a randomized double blind crossover phase where half of the patients are randomized to DBS on, followed by DBS off and the other half starts with DBS off followed by on (Bergfeld et al., 2016b).

## Methods

### Participants

Patients were recruited at two hospitals in the Netherlands, Academic Medical Center in Amsterdam (AMC) and St. Elisabeth Hospital in Tilburg (SEH). They had a primary diagnosis of major depressive disorder (MDD) with illness duration of at least two years. They had to be treatment resistant, defined as an inadequate response to at least two different classes of second generation antidepressants, one tricyclic antidepressant, one tricyclic antidepressant with lithium augmentation, one monoamine oxidase inhibitor and six or more sessions of bilateral electroconvulsive therapy. Symptom severity was assessed using the clinician rated Hamilton rating scale for depression (HDRS)(M. Hamilton, 1960) and Montgomery-Asberg depression rating scale (MADRS)(Montgomery & Asberg, 1979) and the self-rated inventory of depressive symptomology (IDS-SR)(John Rush et al., 1986). The minimum HDRS score for inclusion was higher than 17 and a Global Assessment of Function score had to be lower than 46 (“Diagnostic and Statistical Manual of Mental Disorders, 5th Edition,” 2013). Two electrodes (Model 3389, Medtronic inc.) with four contact points of 1.5 mm, intersected by 0.5 mm were implanted through the internal capsules. The deepest contact was located in the nucleus accumbens (NAc), the three upper contact points were located in the ventral anterior limb of the internal capsule (vALIC).

Patients then underwent an optimization phase where patients were evaluated biweekly and the stimulation parameter adjusted to achieve optimum response. The optimization phase ended either after a four week stable response or when a maximum of 52 weeks was reached. After the optimization the patients underwent a crossover phase were they were randomly assigned to either have DBS on or a week followed by DBS off or vice versa. Symptom severity according to the clinical scales was collected at baseline and at the end of each phase. Twenty healthy controls were recruited for the imaging comparisons. The study was approved by the medical ethics board of both hospitals (Netherlands Trial Register: NTR2118). All patients and controls signed an informed consent form.

### Image acquisition

Each patient underwent a scanning session pre-operatively and a second scanning session after the optimization phase, for comparison of these two states. Each session included a resting state fMRI scan and an anatomical scan. Thereafter, similar scans were obtained with 1-6 weeks between the scanning sessions, after the end of each cross-over phase. The controls were also scanned twice with one year between sessions.

Images were acquired using a 1.5T Siemens MAGNETOM Avanto scanner. A transmit receive head coil was used to minimize exposure of DBS electrodes to the pulsed radiofrequency field. The DBS was turned off a few minutes prior to scanning. The head of the patient was held in place with padding and straps. Specific absorption rate was limited to 0.1 W kg^-1^. Structural images were acquired with 1x1x1 mm resolution using a 3D sagittal MPRAGE with repetition time (TR) of 1.9 s, echo time (TE) of 3.08 ms, flip angle of 15° and inversion time (TI) of 1.1 s. FMRI data were acquired with two dimensional echoplanar imaging with TR = 2000 ms, TE = 30 ms and a flip angle = 90°. Each scan consisted of 25 transversal slices of 4 mm with voxel size of 3.6x3.6 mm and slice gap of 0.4 mm. The first ten volumes were discarded to allow for magnetization stabilization and the subsequent 180 volumes were analyzed. Total scanning time was 360 seconds resulting in 180 3d volumes.

### Image preprocessing

Image processing was performed as described previously (Fridgeirsson et al., 2020b). The functional data were realigned to the first volume. Then the structural data were brain extracted and coregistered with the functional data. Then the data were normalized to MNI (Montreal Neurological Institute) using the unified segmentation (Ashburner & Friston, 2005) approach in SPM12 (http://www.fil.ion.ucl.ac.uk/spm) which has been shown to be robust to brain lesions (Crinion et al., 2007). The functional data were then resampled to 2mm isotropic, smoothed with an 8mm Gaussian kernel and bandpass filtered between 0.01 – 0.1 Hz.

### Functional connectivity analysis

We performed seed-based functional connectivity (FC) analyses for the laterobasal (LB) amygdala and NAc as these were the regions where previous functional connectivity changes had been found using the same target in OCD (Figee et al., 2013b; Fridgeirsson et al., 2020b). For the LB amygdala the analysis followed that of Fridgeirsson et al (Fridgeirsson et al., 2020a). For the NAc it did as well except the probability map used was from the subcortical atlas from (Pauli et al., 2018).

Since the dropout region due to the electrode artifacts intersects with the NAc we removed it from the seed region. We subtracted the post-operative normalized mean functional fMRI from the pre-operative scan, which had no dropout. For all subsequent analyses the dropout regions were removed from the NAc. To further check the effect of this step we calculated the volume of the removed region and correlations between timeseries from seed regions in pre-operative scans including and excluding the dropout. The average proportion of removed volume from the NAc due to dropout was 22% and 20% for the left and right NAc. The average correlation between the NAc and NAc-without–dropout timeseries was 0.84 and 0.89 for the left and right NAc.

To estimate that the effects of motion have been accounted for the quality control benchmarks from Parkes et al (Parkes et al., 2018) were used using the parcellation from Gordon et al (Gordon et al., 2016). The proportion of significant correlations between framewise displacement and FC was 0.05. The median correlation was 0.11 and the Spearman correlation between the FC and Euclidean distance was - 0.06. Comparing to the figures in (Parkes et al., 2018), this suggest our preprocessing strategy is accounting well for the effects of motion.

For the pre vs post comparison the statistical maps were entered into a 2x2 factorial design in SPM12. The factors were Group (patient vs controls) and Time (pre-operative vs after optimization). Voxel-wise statistical tests were corrected for family wise error at the cluster level (p<0.05) with a threshold of p=0.001 (Eklund et al., 2016), across the entire brain or within a priori regions of interest (Fridgeirsson et al., 2020b). Post hoc tests were performed for significant interactions. Same procedure was used for the crossover comparison but with paired t-tests.

### Effective connectivity analysis

To investigate directionality of the connections for the network we used spectral DCM (K. J. Friston et al., 2014b). The same procedure as in Fridgeirsson et al (Fridgeirsson et al., 2020a) was followed to build DCM models with four ROIs (left LB amygdala, left NAc, left Insula and vmPFC) as nodes and bilateral connections between all regions defined, resulting in 16 connections including each region’s self-connection.

After individual models were inverted a second level parametric empirical Bayes (PEB) model was constructed to model the effect of between subject effects on the estimated effective connections (K. J. Friston et al., 2016; Zeidman et al., 2019). After estimating the group-level PEB model we performed a search over nested PEB models by pruning those parameters that did not contribute to the model evidence(K. Friston & Penny, 2011; Rosa et al., 2012). Bayesian model averaging was then used after the final iteration to determine the strength of connections in the last Occam’s window of 256 models. Free energy of models with vs without each connection were used to compute the posterior probabilities (PP) for each connection. Connections were included in the results if they had very strong amount of evidence (PP > 0.99). PEB is a multivariate Bayesian GLM in which all model parameters are fit at once and no multiple comparisons correction is required. For the pre-operative vs after optimization comparison the modelled effect was the interaction of group (Patients vs Controls) and time (Pre-operative vs after-optimization). For the crossover phase it was the effect of DBS on vs off.

## Results

In thirteen of the 25 patients from our study on DBS in depression (Bergfeld et al., 2016b) complete imaging sets at baseline and after optimization were available. Three patients were excluded due to excessive head motion and in one patient dicom files were corrupt, resulting in a sample of 9 patients for the pre-surgical to after-optimization comparison. Twenty healthy controls were recruited, of whom three did not complete two scanning sessions. In one the dicom files were corrupted and four were excluded due to excessive head motion during scanning, resulting in a sample of 12 controls. The patients and controls did not differ in age, sex, education level or head motion during scanning (table 1). The optimization phase resulted in significant reduction in MADRS scores. Comparable changes in HAM-D and IDS scores were observed, but did not reach significance. Changes in MADRS and HAM-D were highly correlated during optimization (r=0.91) and crossover (r=0.99) while MADRS/HAM-D were less correlated to IDS during optimization (r=0.87/0.78) than during crossover (r=0.97/0.98). For the crossover phase turning DBS off resulted in significantly worse clinical scores overall (table 1).

**Table 1:** Demographics of study sample and clinical scales for pre-operative vs after optimization.

| Demographics long term comparison | Patients (n=9) |  | Controls (n=12) |  | Difference |
| --- | --- | --- | --- | --- | --- |
|  | Mean (SD) | Range | Mean (SD) | Range | P-value |
| Age, y. | 52.6 (5.5) | 47-62 | 53.3 (8.1) | 31-62 | 0.83<sup>1</sup> |
| Sex (n, % Males). | n=2, 22.2% |  | n=5, 41.7% |  | 0.096<sup>2</sup> |
| Education level. § | 4.2 (2.0) | 2-7 | 3.6 (1.7) | 2-7 | 0.44<sup>1</sup> |
| Illness Duration, y. | 16.6 (8.3) | 3-31 |  |  |  |
| Motion (mean framewise displacement), mm. | 0.22 (0.098) | 0.09-0.45 | 0.19 (0.049) | 0.09-0.30 | 0.15<sup>1</sup> |
| Clinical scales long term comparison | Pre-operative |  | After optimization |  | P-value |
|  | Mean (SD) | Range | Mean (SD) | Range |  |
| HAM-D | 23.3 (6.6) | 16-33 | 15.3 (9.5) | 4-34 | 0.06<sup>3</sup> |
| MADRS | 34.4 (7.8) | 23-44 | 22.6 (13.3) | 5-44 | 0.016<sup>3*</sup> |
| IDS | 49.7 (7.8) | 36-61 | 38.0 (16.7) | 13-71 | 0.045<sup>3</sup> |
| Clinical scales crossover patients (n=9) | DBS on |  | DBS off |  | P-value<sup>3</sup> |
|  | Mean (SD) | Range | Mean (SD) | Range |  |
| HAM-D | 14.3 (7.3) | 5-28 | 22.4 (5.1) | 14-29 | 0.009* |
| MADRS | 21.9 (12.4) | 6-44 | 34.3 (7.4) | 21-44 | 0.012* |
| IDS | 32.2 (16.4) | 10-59 | 45.9 (9.5) | 32-58 | 0.003* |

Abbreviations: HAM-D: Hamilton Rating Scale for Depression; MADRS: Montgomery-Asberg Depression Rating Scale, IDS: Inventory for Depression Symptomology
§ According to the International Standard Classification of Education (ISCED) 2011
\* $p < 0.05$ corrected for three comparisons. <sup>1</sup>independent sample t-test. <sup>2</sup>Chi-square test. <sup>3</sup>Paired t-test

## Functional connectivity

### Pre-operative vs post-optimization

Left LB amygdala connectivity showed a significant interaction between group and time with the left precentral cortex (MNI: (−10, -30, 44), size: 904 mm^3^, p=0.001, FWE-corrected, Figure 1). According to the Harvard Oxford cortical atlas the cluster is 23% in the posterior cingulate cortex and 64% in the precentral gyrus (Makris et al., 2006). Post hoc tests revealed significant decrease of LB amygdala and precentral cortex connectivity in controls over time (p=0.007), whereas it tended to increase (non-significantly) after DBS treatment in patients. The results presented in Figure 1 suggest that there was one outlier, though the results remained significant after excluding this participant. The group by time interaction did not reveal any significant clusters for the right LB amygdala as seed.

**Figure 1:**
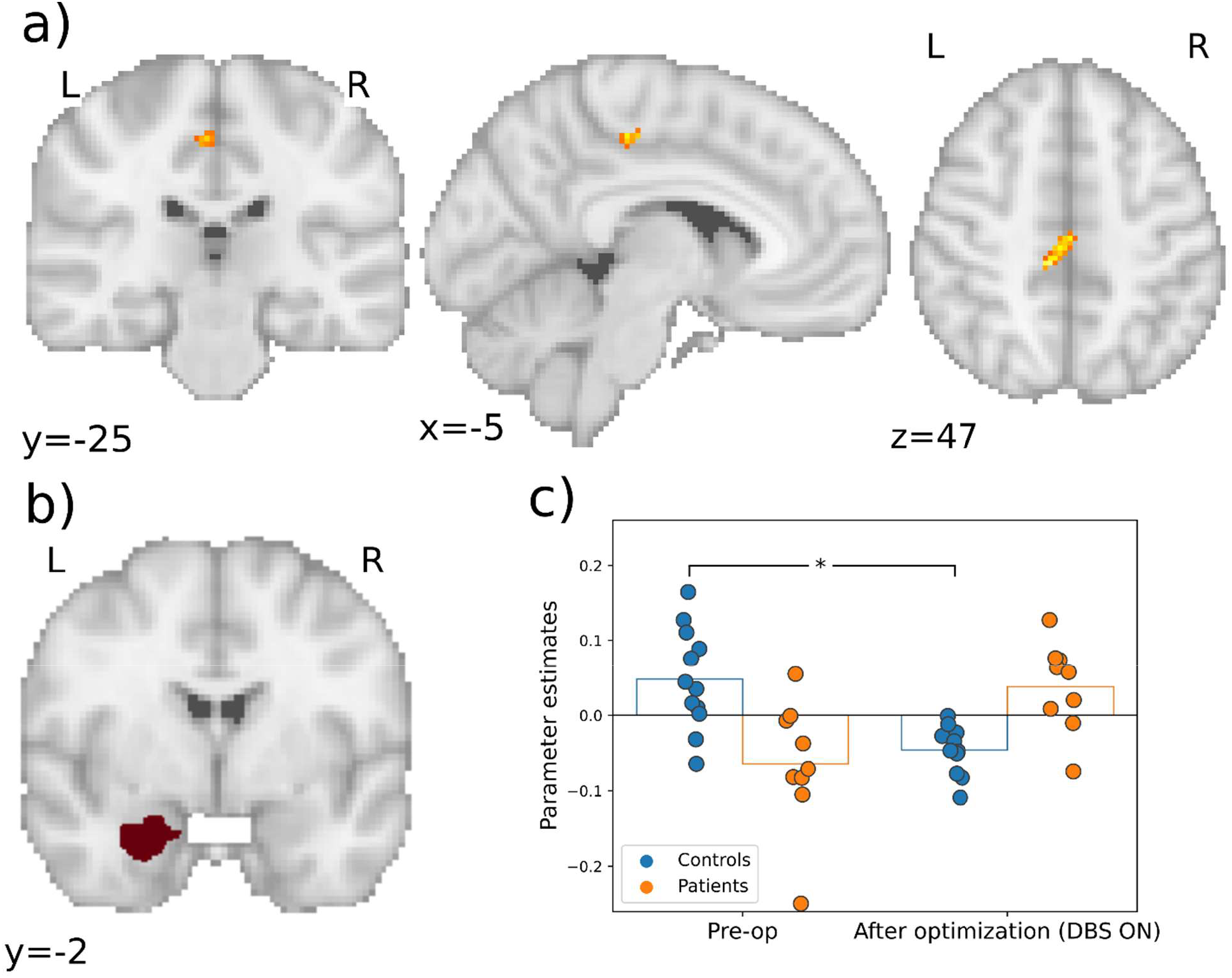
Whole brain significant connectivity changes between the left LB amygdala and precentral cortex, a) coronal, sagittal and axial views of the significant cluster b) The seed region in the left LB amygdala c) The parameter estimates showing the significance from post-hoc testing. *p<0.05.

In addition, there was a significant interaction between group and time with left LB amygdala connectivity anterior in the left insula (MNI: (−32, 26, 6), size: 136 mm^3^, p=0.027, FWE with small volume(SV) correction; Figure 2). Post hoc tests with SV correction revealed a significant decrease in controls from the first to the second session (p=0.035). DBS tended to increase the connectivity in patients from before surgery to after optimization.

**Figure 2:**
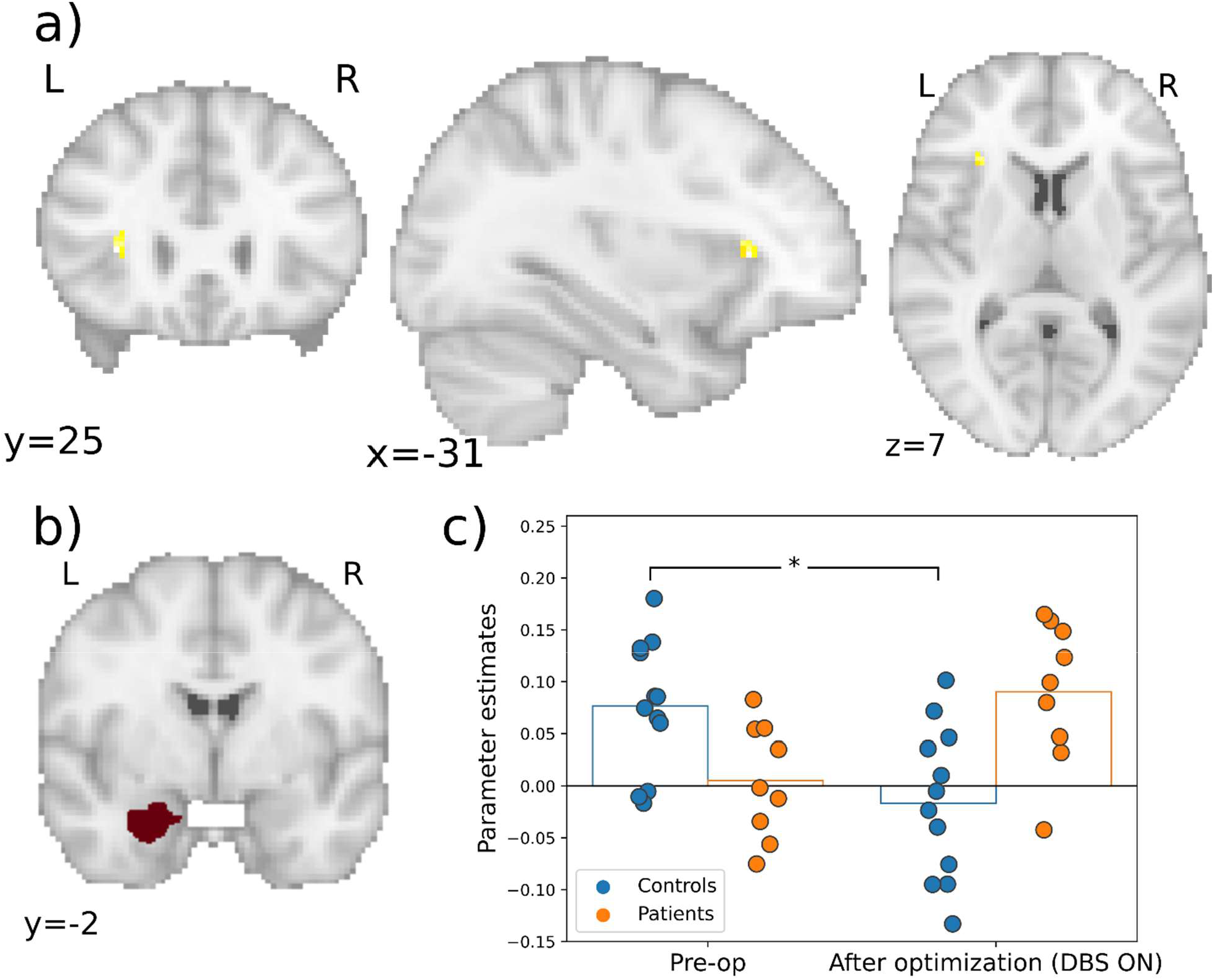
Significant connectivity changes between the left LB amygdala and left insula. a) Cluster in left anterior insula. b) The seed region in the left LB amygdala c) Parameter estimates with significance from post hoc testing.

Left NAc connectivity showed a significant interaction between group and time (MNI: (4, 58, 0), size: 176 mm^3^ p=0.013, FWE-correction) with the most anterior part of the right vmPFC. Connectivity tended to decrease in patients from before surgery to after optimization while it increased over time in controls, though these post-hoc tests were not significant (Figure 3).

**Figure 3:**
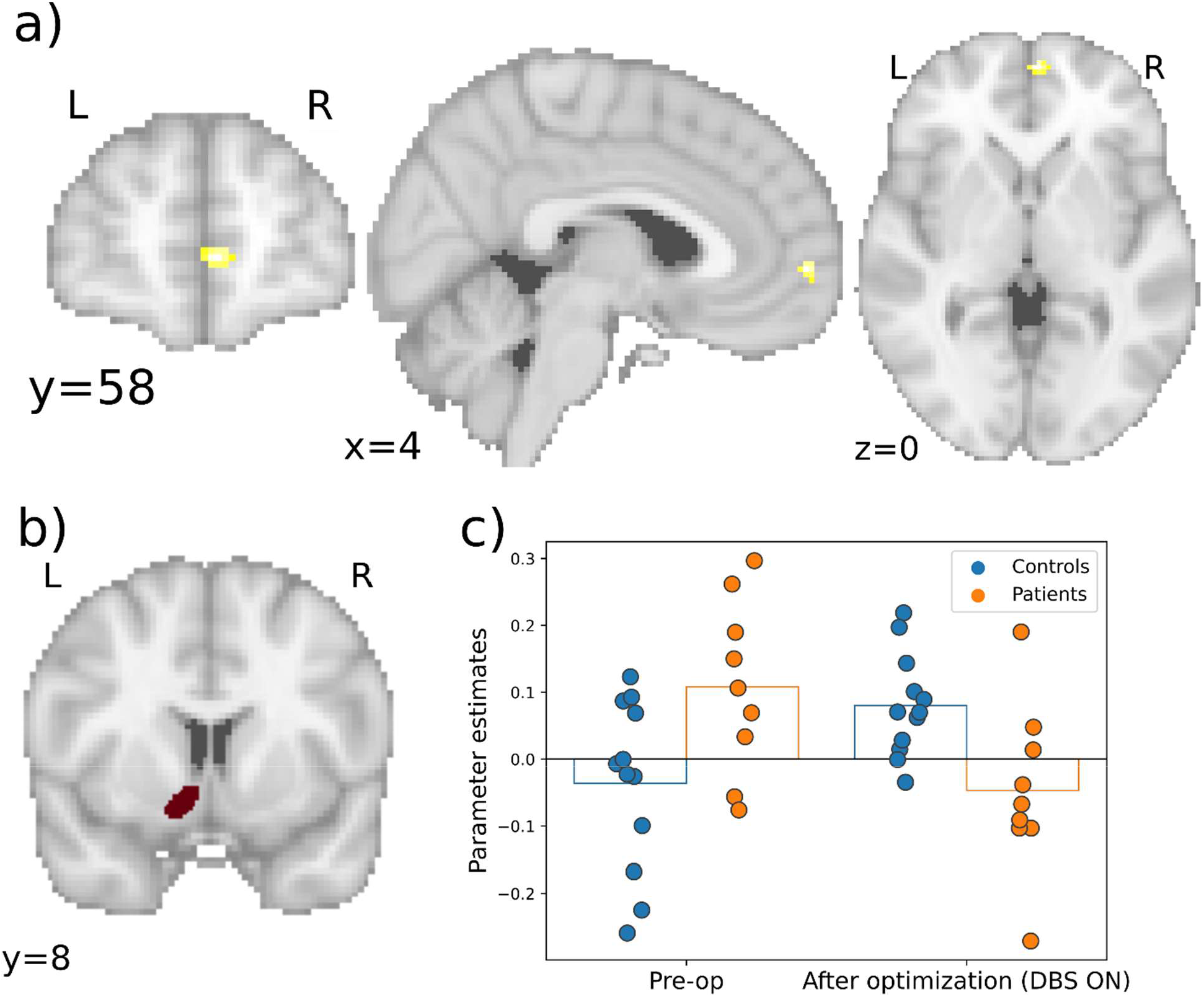
Significant connectivity changes between left NAc and vmPFC. a) coronal, sagittal and axial views of the significant cluster in the vmPFC. b) The seed region in left NAc. c) Parameter estimates.

Right NAc connectivity showed a significant interaction between group and time with the posterior, almost subgenual part of vmPFC (MNI: (6, 36, -10), size: 496 mm^3^, p=0.044, FWE-corrected). Post hoc tests revealed a significant decrease in connectivity with this cluster in patients from before surgery to after optimization (p=0.001). Further this connectivity was significantly lower in patients than controls after optimization (p<0.001; Figure 4).

**Figure 4:**
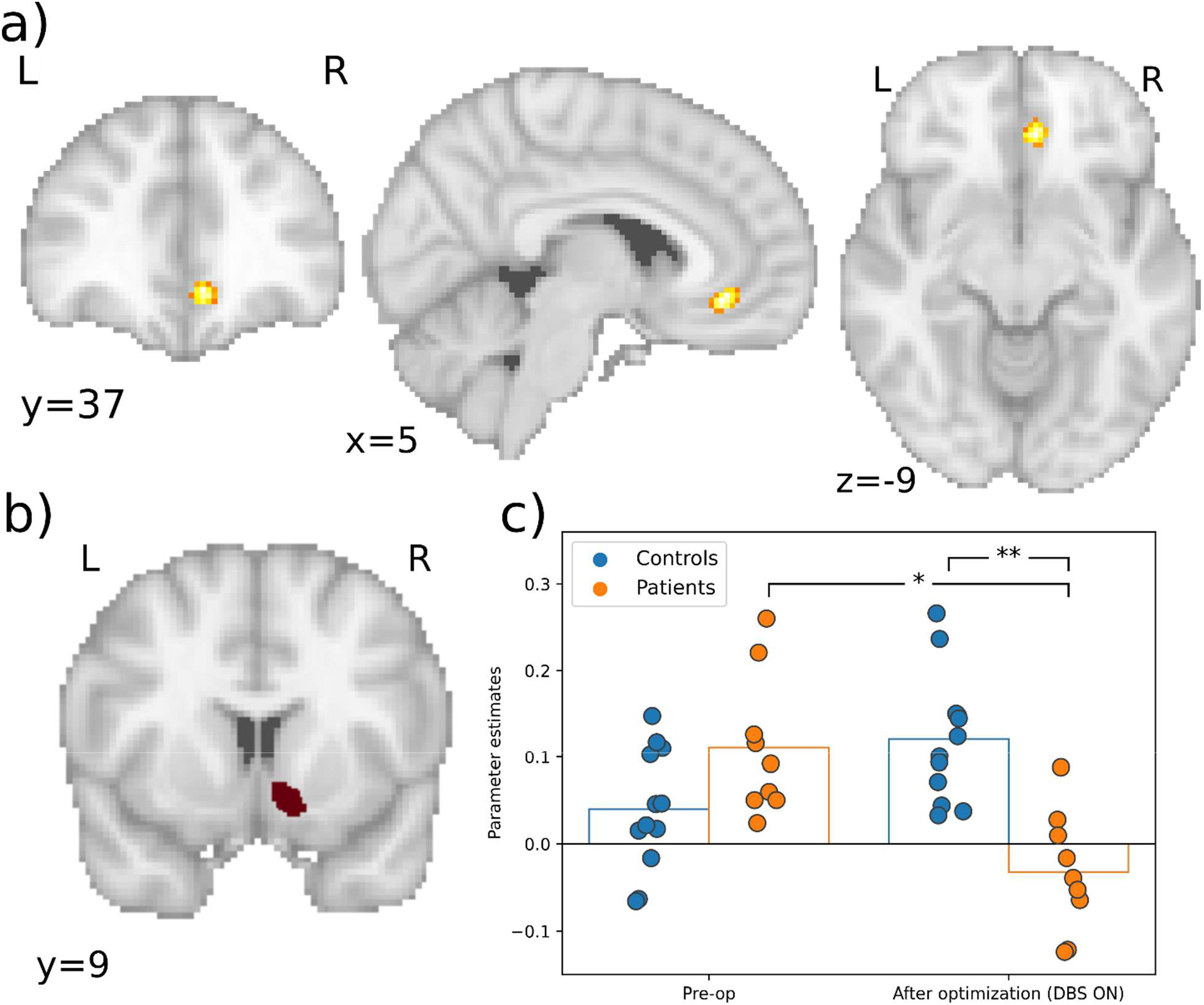
Significant connectivity changes between right NAc and vmPFC. a) coronal, sagittal and axial views of the significant cluster in the vmPFC. b) The seed region in the right NAc. c) Parameter estimates with significance from post hoc testing.

### Crossover phase

For the crossover phase functional connectivity changes with neither the LB amygdala nor NAc were significant.

## Effective connectivity

### Pre-operative vs post-optimization

The average explained variance of the fitted individual models was 84% with a standard deviation of 4.9%. The results of the PEB model can be seen in Figure 5a and table 3. The interaction between group and time demonstrated changes in network connections in which optimized DBS-treatment was associated with weaker self-inhibition of the vmPFC and weaker excitatory connection from vmPFC to amygdala (−0.11 Hz) in patients compared to controls. After DBS-therapy there was also less inhibitory connection from amygdala to insula (0.09 Hz), with weaker excitatory connection from insula to amygdala (−0.09 Hz), and greater inhibitory connection from NAc to amygdala (−0.11 Hz) in patients compared to controls.

**Figure 5:**
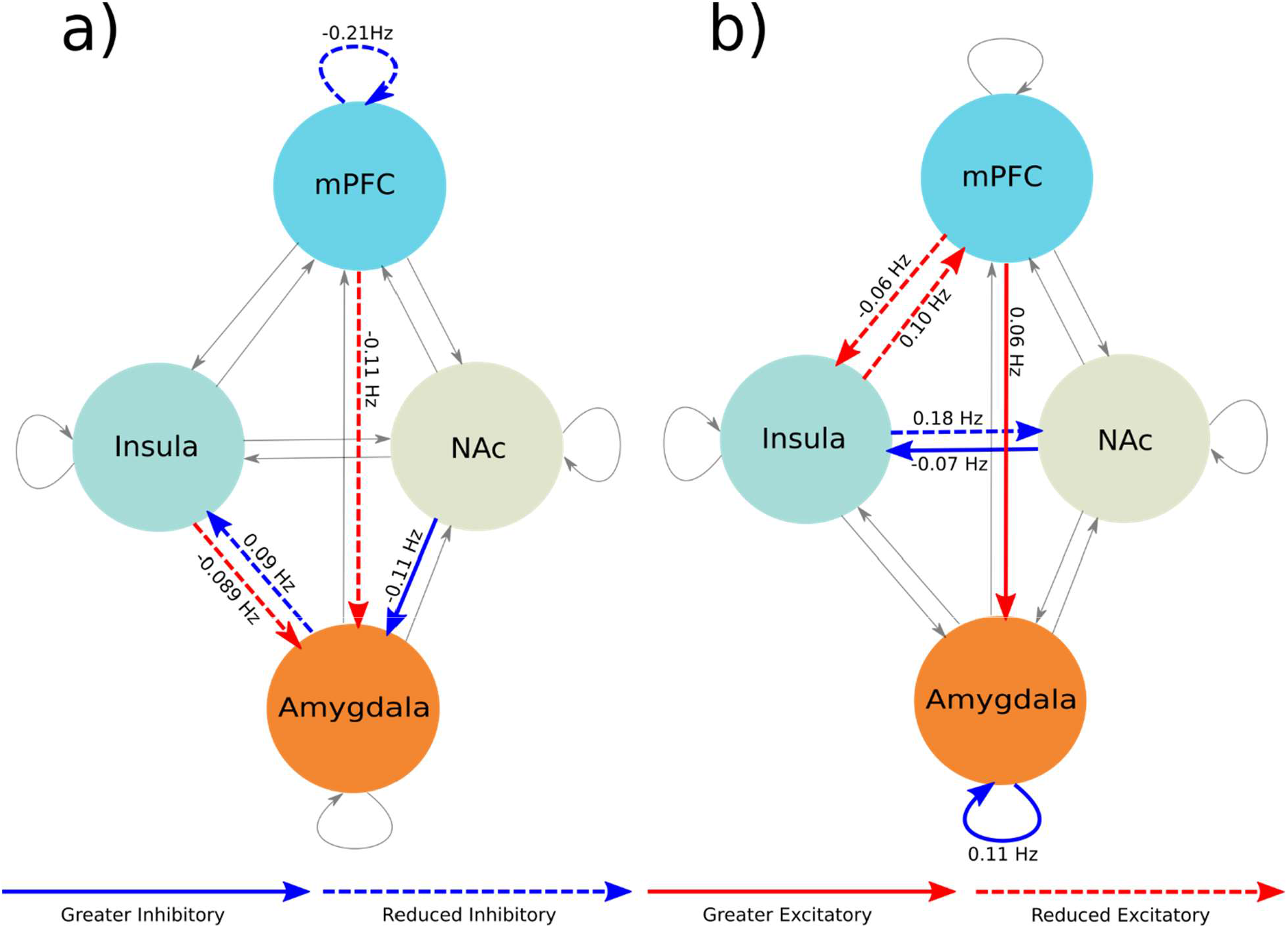
Changes in excitatory and inhibitory connections a) After DBS treatment vs pre-surgical network changes compared to changes in controls. b) Crossover phase network changes associated with DBS on in patients. Grey connections were not significant.

**Table 3:** Parameter estimates for effective connectivity.

**Pre vs post optimization**
| **Connection** | **Parameter estimates**<br>**(90% credible interval)** | **Average**<br>**connectivity** | **Direction of**<br>**changes** | **Posterior**<br>**probability** |
| --- | --- | --- | --- | --- |
| **NAC → Insula** | **-0.11 Hz (-0.057 to -**<br>**0.16)** | **Inhibitory** | **Greater inhibitory** | **1.0** |
| **Insula → Amygdala** | **-0.089 Hz (-0.035 to -**<br>**0.14)** | **Excitatory** | **Reduced excitatory** | **1.0** |
| **vmPFC → Amygdala** | **-0.11 Hz (-0.064 to -**<br>**0.16)** | **Excitatory** | **Reduced excitatory** | **1.0** |
| **Amygdala → Insula** | **0.09 Hz (0.024 to 0.156)** | **Inhibitory** | **Reduced inhibitory** | **1.0** |
| **vmPFC self-inhibition** | **-0.21 (-0.29 to -0.15)** | **Inhibitory** | **Reduced inhibitory** | **1.0** |
| **Crossover phase** |  |  |  |  |
| **Amygdala self-**<br>**inhibition** | **0.11 Hz (0.017 to**<br>**0.21)–** | **Inhibitory** | **Greater inhibitory** | **1.0** |
| **vmPFC → amygdala** | **0.06 Hz (0.027 to 0.010)** | **Excitatory** | **Greater excitatory** | **1.0** |
| **Insula → NAc** | **0.18 Hz (0.13 to 0.24)** | **Inhibitory** | **Reduced inhibitory** | **1.0** |
| **NAc → insula** | **-0.07 Hz (-0.018 to -**<br>**0.12)** | **Inhibitory** | **Greater inhibitory** | **1.0** |
| **vmPFC → insula** | **-0.06 Hz (-0.024 to -**<br>**0.098)** | **Excitatory** | **Reduced excitatory** | **1.0** |
| **Insula → vmPFC** | **0.10 Hz (0.055 to 0.14)** | **Excitatory** | **Reduced excitatory** | **1.0** |

### Crossover phase

Average explained variance was 83.7% with a standard deviation of 4.4%. The results for the PEB model for the crossover phase can be seen in Figure 5b and table 3. After the crossover phase with DBS on, there was greater excitatory connection from the vmPFC to the amygdala (0.06 Hz) and with stronger self-inhibition in the amygdala (0.11 Hz). DBS on was also associated with reduced excitatory connection from vmPFC to insula (−0.06 Hz) and from insula to vmPFC (0.10 Hz), as well as with weaker inhibitory connection from insula to NAc (0.18 Hz) and stronger inhibitory connection the NAc to insula (−0.07 Hz).

## Discussion

We investigated the neural mechanism underlying the effects of vALIC DBS in MDD. DBS treatment in patients increased resting-state connectivity of the left LB amygdala with the precentral cortex and anterior part of the left insula from before operation to after optimization in comparison to changes in connectivity in controls. In addition, DBS decreased connectivity between the left and right NAc with the the right vmPFC. Effective connectivity modeling using spectral DCM revealed widespread amygdala-centric changes from before operation to after optimization in patients compared to controls. Whereas short term changes in effective connectivity during the blind crossover phase primarily revealed reduced insular excitatory connectivity with vmPFC and net insular inhibition from NAc in active compared to sham stimulation.

Once DBS treatment was optimized, functional connectivity between left LB amygdala and left insula had increased while it decreased between bilateral NAc and vmPFC. As we previously investigated the effects of vALIC DBS in OCD, this enables us to interpret these results in the context of the results in OCD. However, it is important to consider that we used a different design in which the DBS device was turned off for approximately one week after one year of active stimulation. In OCD, we observed that vALIC DBS led to comparable changes in NAc-vmPFC connectivity (Figee et al., 2013b) but to opposite changes in LB amygdala-insula connectivity (Fridgeirsson et al., 2020b). This suggests that vALIC DBS induces generic effects that are independent of the underlying pathophysiology, as well as neural effects that are disorder specific. Frontostriatal hyperconnectivity has been considered core part of the pathology of OCD (Pauls et al., 2014), whereas multiple studies have suggested that functional connectivity between NAc and vmPFC is reduced in depressed patients (Furman et al., 2011; Liu et al., 2021a; Satterthwaite et al., 2015; Zhou et al., 2022). However, more recent large-scale multicenter studies showed that resting-state connectivity is reduced throughout the brain in both MDD as well as OCD, with the exception of primarily thalamic hyperconnectivity(Bruin et al., 2023; Gallo et al., 2023; Javaheripour et al., 2021). The pathophysiology of MDD and OCD may therefore be more comparable than previously thought. It is important to consider that these studies have largely been conducted in non-refractory patients, and NAc connectivity may be different in patients indicated for DBS. Regardless, our current results indicate that the effect of vALIC DBS on frontostriatal connectivity, now observed in two disorders, could thus be a generic effect of the DBS treatment, which may relate to the implantation of the electrodes close to the NAc.

In OCD, we found decreased LB amygdala - insula connectivity with DBS treatment which was associated with mood and anxiety changes (Fridgeirsson et al., 2020b). Here we found the opposite effect in MDD, where DBS increased connectivity between these regions in depression. In non-refractory depression, differences in amygdala connectivity between patients and controls has suggested reduced connectivity in patients (Ramasubbu et al., 2014; Tang et al., 2018). Considering these opposite effects of DBS in the same target across the two disorders, the effects of DBS on this connectivity seem to be dependent on the underlying pathophysiology. It should also be pointed out that the effect in insular connectivity was measured in the most anterior part of the insular cortex in MDD, whereas in OCD these effects were seen in the posterior area of the insula, suggesting a differential functional organization within the insula. Further, we did find changes in connectivity between LB amygdala and medial precentral gyrus where connectivity increased with DBS but no such changes were detected in the previous OCD study.

The regions implicated here with the treatment effect of vALIC DBS in MDD have various roles in the hypothesized mechanism of MDD from the literature. The vmPFC is a part of the default mode network which is commonly associated with increased rumination in the pathophysiology of MDD (Sheline et al., 2009). The NAc on the other hand is a part of the reward network. But as decreased functional connectivity between those regions has been implicated with anhedonia in MDD (Liu et al., 2021b), the DBS related decrease in NAc-vmPFC connectivity and improvement of anhedonia appears paradoxical. However, vmPFC functioning is altered in refractory MDD compared to non-refractory MDD (Runia et al., 2022). We therefore speculate that this could be due to altered NAc-vmPFC connectivity in refractory MDD as well.

There were marked differences between the long term (preoperative vs post-optimization) and the short term (crossover phase) results. For functional connectivity, all detected changes were for the long term comparison while for effective connectivity there were both long term and short term changes. The long term changes were amygdala centric while the short term changes were insula centric. This could indicate that during the long optimization phase rebalancing of connectivity between the nodes takes place. Then when entering the crossover phase this balance is perturbed in a way that is different from the pre-operative baseline. These short term changes are as well not detected by functional connectivity, only with the spectral DCM. While functional connectivity only measures covariance at zero-lag using the blood oxygen level dependent response, spectral DCM fits the whole cross-covariance and cross-spectrum of the time series, including an model for how the neural activity results in hemodynamic changes. Further spectral DCM does apply smoothness constraints by using an autoregressive model to fit the spectrum which should reduce overfitting. Taken together this indicates spectral DCM is a more general connectivity metric that takes more information into account than functional connectivity and our results indicate it can even be more sensitive to network changes due to an intervention such as DBS.

The strengths of the study are that we for the first time report the effects of vALIC DBS for depression on resting-state connectivity on the short and long term. We further limit ourselves to exploring regions that have been implicated using DBS of the same target in OCD (Figee et al., 2013b; Fridgeirsson et al., 2020b). Further for the long term comparison a particular strength is the inclusion of a control group that allows us to account for repeated testing. There are some limitations to our study as well. First of all, we have a small sample, which unfortunately cannot be prevented given the small number of MDD patients treated with DBS electrodes. However, to our knowledge this the largest sample using functional neuroimaging in DBS for MDD. This study was part of a randomized controlled clinical trial which was determined to have enough power to detect clinical effects of vALIC DBS in MDD (Bergfeld et al., 2016b). The imaging outcomes were secondary outcomes and unfortunately complete fMRI data was missing for more than half of patients. The small sample did not allow for investigating heterogeneity or effects of other factors such as medication use as well as limiting us to detect large effects. Second, because of computational reasons we cannot include all nodes of interest in our effective connectivity analysis. This could mean that some of the changes we report are caused or mediated by unmodelled nodes.

In conclusion, we found that vALIC DBS treatment in MDD patients modulates resting-state connectivity in a limbic network. Some of these changes appear to be a generic effect of vALIC DBS while other appear to be disorder specific. Using effective connectivity we found long-term changes centered around the amygdala and shorter-term changes which are more insula centric. This is an indication of complex rebalancing of interceptive and emotional processing changes induced by DBS treatment which can inform future research to improve DBS treatment for MDD.

## Data Availability

All data produced in the present study are available upon reasonable request to the authors

## Notes

### Competing Interest Statement

The authors declare the following financial interests/personal relationships which may be considered as potential competing interests: This investigator-initiated study was funded by Medtronic Inc (25 DBS systems, in kind). The funders had no role in the design, execution, and analysis of the study, nor in writing of the manuscript or the decision to publish. Isidoor Bergfeld, Pepijn van den Munckhof, Rick Schuurman, Damiaan Denys, and Guido van Wingen currently execute an investigator-initiated clinical trial on deep brain stimulation for depression, which is funded by Boston Scientific (24 DBS systems in kind) and a grant of ZonMw (nr. 636310016). Rick Schuurman acts as consultant for Boston Scientific and Medtronic on educational events. All other authors do not declare any conflicts of interest.

### Clinical Trial

NTR2118

### Funding Statement

This investigator-initiated study was funded by Medtronic Inc (25 DBS systems, in kind).

### Author Declarations

The ethics committees of the Academic Medical Centre, Amsterdam (AMC) and St Elisabeth Hospital, Tilburg (SEH) gave ethical approval for this work.

